# Tracing the trajectories of SARS-CoV-2 variants of concern between December 2020 and September 2021 in the Canary Islands (Spain)

**DOI:** 10.1101/2022.02.26.22271544

**Authors:** Laura Ciuffreda, Rafaela González-Montelongo, Julia Alcoba-Florez, Diego García-Martínez de Artola, Helena Gil-Campesino, Héctor Rodríguez-Pérez, Antonio Íñigo-Campos, Isabel De Miguel-Martínez, Tomás Tosco-Nuñez, Oscar Díez-Gil, Agustín Valenzuela-Fernández, José M. Lorenzo-Salazar, Carlos Flores

## Abstract

Several variants of concern (VOCs) explain most of the Severe Acute Respiratory Syndrome Coronavirus 2 (SARS-CoV-2) epidemic waves in Europe. We aimed to dissect the spread of the SARS-CoV-2 VOCs in the Canary Islands (Spain) between December 2020 and September 2021 at a micro-geographical level. We sequenced the viral genome of 8,224 respiratory samples collected in the archipelago. We observed that Alpha (B.1.1.7) and Delta (B.1.617.2 and sub-lineages) were ubiquitously present in the islands, while Beta (B.1.351) and Gamma (P.1/P.1.1) had a heterogeneous distribution and were responsible for fewer and more controlled outbreaks. This work represents the largest effort for viral genomic surveillance in the Canary Islands so far, helping the public health bodies in decision-making throughout the pandemic.

## Introduction

The continuous emergence and international widespread of variants of concern (VOCs) of the Severe Acute Respiratory Syndrome Coronavirus 2 (SARS-CoV-2) have revealed the occurrence of multiple viral spike gene mutations that promote increased transmissibility and some degree of immune escape (Tao *et al*., 2021). In response, the public health system activated the organisation of centres for routine viral genome surveillance starting in mid-January 2021 in our laboratories, which were later distinguished as the Reference Centre for the Network of COVID-19 Genomic Surveillance in the Canary Islands.

We have previously tracked the entrance and surge of one of the three most widely distributed VOCs, B.1.1.7 (Pango lineage nomenclature (Rambaut *et al*., 2020)), or Alpha variant (WHO, 2022), in December 2020 in Tenerife (Alcoba-Florez *et al*., 2021) due to the island direct flight connections with the UK where it may have originated. This lineage has been associated with increased transmissibility (Davies *et al*., 2021; Volz *et al*., 2021) compared to the previously circulating variants, a feature that has been linked to the presence of the mutation N501Y in the Spike (S) receptor-binding domain (RBD). Other VOCs that have also emerged in late 2020 are B.1.351, or the Beta variant, and P.1/P.1.1, or the Gamma variant, firstly identified in South Africa (Tegally *et al*., 2021) and in Brazil (Faria *et al*., 2021), respectively. These lineages have been associated with increased transmission (Faria *et al*., 2021; Pearson *et al*., 2021; Tegally *et al*., 2021) and carry multiple mutations affecting the RBD, most importantly the N501Y (such as Alpha), and K417N/K417T and E484K which reduce the effectiveness of some vaccines (Garcia-Beltran *et al*., 2021) and specific monoclonal antibody treatments (Greaney *et al*., 2021; Harvey *et al*., 2021). Later in time, lineage B.1.617.2, or the Delta variant, was identified in India where it was responsible for a surge of COVID-19 cases in April 2021 (Dhar *et al*., 2021). The Delta variant presented additional mutations, such as the L452R in the RBD and the P618R, and has been characterized by higher transmissibility (F. Campbell *et al*., 2021) and reduced sensitivity to antibody neutralization (Kuzmina *et al*., 2021; Liu *et al*., 2021; Planas *et al*., 2021; Wall *et al*., 2021) and vaccine effectiveness (Farinholt *et al*., 2021; Kuzmina *et al*., 2021; Lopez Bernal *et al*., 2021; Planas *et al*., 2021). From the Delta variant, numerous sub-lineages have emerged, reporting the presence of additional mutations such as the Y145H and the Y222V in the N-terminal domain of the S protein found in the AY.4.2 lineage (Mullen *et al*., 2020), which was suggested to be more transmissible than the original variant (Public Health England, 2021). Delta and its sub-lineages became prevalent and completely displaced Alpha worldwide throughout the summer of 2021 (WHO, 2022).

Here, we describe the introduction and temporal evolution of these four VOCs in the Canary Islands (Spain). The archipelago is formed by eight islands, four occidental (Tenerife, La Palma, La Gomera, and El Hierro) belonging to the Tenerife province, and four oriental islands (Gran Canaria, Fuerteventura, Lanzarote, and La Graciosa) belonging to the Gran Canaria province. The relative geographical isolation of the archipelago and its islands, and their central role as a hub for international tourism and in the European migratory crisis have shaped the COVID-19 pandemic and delineated the introduction and spread of these VOCs in the territory. In fact, the first SARS-CoV-2 outbreaks in Spain were detected in La Gomera and Tenerife islands (January and February 2020, respectively; available reports from: https://www.isciii.es/QueHacemos/Servicios/VigilanciaSaludPublicaRENAVE/EnfermedadesTransmisibles/Documents/INFORMES/Informes%20COVID-19/Informe%20COVID-19.%20N%c2%ba%201_11febrero2020_ISCIII.pdf).

## Materials and methods

### Design and sample collection

The study was conducted at the University Hospital Nuestra Señora de Candelaria (HUNSC) (Santa Cruz de Tenerife, Spain). The institutional review board approved the study (approval number: CHUNSC_2020_24). We assessed nasopharyngeal swabs from COVID-19/SARS-CoV-2 patients from the 18^th^ of December 2020 to the 27th of September 2021. Routine COVID-19 testing in the centres was conducted using diverse commercial RT-qPCR alternatives, as described elsewhere (Alcoba-Florez *et al*., 2020). In line with the guidelines indicated by the ECDC (European Centre for Disease Prevention and Control, 2021) and assuming a prevalence of 10,000 positive cases per month in the territory (https://opendata.sitcan.es/dataset/datos-epidemiologicos-covid-19), we estimated the sequencing of 12,000 samples a year for accurate tracking of viral variants (at 50% accuracy in prevalence estimates).

### Viral genome sequencing and classification

Samples were selected for sequencing if they showed a cycle threshold (Ct) ≤30 for any of the amplicon targets included in the COVID-19 diagnostic kits. Libraries were prepared following either the Midnight protocol v.4 (Freed *et al*., 2020), by means of the Rapid Barcoding kit (SQK-RBK004, Oxford Nanopore Technologies) and sequenced on a MinION and an R9.4 flow cell (Oxford Nanopore Technologies) for 6 hours, or the COVIDSeq Test (Illumina, Inc.) protocol based on the ARTIC V3 amplicons and sequenced on a NextSeq 550 (Illumina) instrument on High Output mode with 36 bp single- or pair-end (after the 7th of September 2021) reads following the procedures described elsewhere (Alcoba-Florez *et al*., 2021; Ciuffreda *et al*., 2021). Positive and negative amplification controls were included in each run (one for each fraction of 94 samples).

The RAMPART (v.1.2.0) software package was used for real-time monitoring of the MinION sequencing run. Reads were basecalled and demultiplexed with Guppy 4.2.2 (high-accuracy mode), and the ARTIC Network bioinformatics procedures (https://github.com/artic-network/artic-ncov2019) were used for read filtering (by length, 250-1500 bp), consensus assembly, and variant calling (nanopolish workflow with maximum coverage of 200X). The COVIDSeq Test reads were processed as described elsewhere (Alcoba-Florez *et al*., 2021) based on the DRAGEN COVIDSeq Test v1.2.2 pipeline and the DRAGEN Lineage v3.5.3 (Illumina, Inc.). Nextclade v.0.14.3 (Aksamentov *et al*., 2021) was used for variant calling and functional predictions. In the final analysis, the sequences having a genome coverage less than 70% of the entire SARS-CoV-2 sequence and having a QC classified as “bad” by Nextclade software were excluded. Pangolin v3.1.1 (O’Toole *et al*., 2021) was used for the classification of the consensus sequences. Microbetrace v0.8.2 (E. M. Campbell *et al*., 2021) was used for cluster investigation together with epidemiological data from the Public Health authorities.

## Results

An overview of COVID-19 cases and sequenced samples in the Canary Islands throughout the study period is presented in **Figure 1A and S1**. Of the 70,588 positive samples collected in the archipelago from the 18^th^ of December 2020 to the 27^th^ of September 2021, 11,956 (16.9%) were sequenced using either Illumina sequencing (11,936) or Oxford Nanopore Technology (20). Of these, 8,224 samples passed the QC filtering steps and had an assigned lineage. In the period, 3,447 samples (41.9%) were assigned to Alpha, 138 (1.7%) to Beta, 47 (0.6%) to Gamma, and 3,262 (39.7%) to Delta and sub-lineages of Delta (1,066 (12.9%)), while 1,330 (16.1%) were assigned to other non-VOC lineages (**Figure 1B**). Within the Delta sub-lineages, AY.4 (153) and AY.12 (119) were the most commonly found throughout the period. None of the Delta sub-lineages was assigned to AY.4.2 (also known as Delta plus).

**Figure 1.**
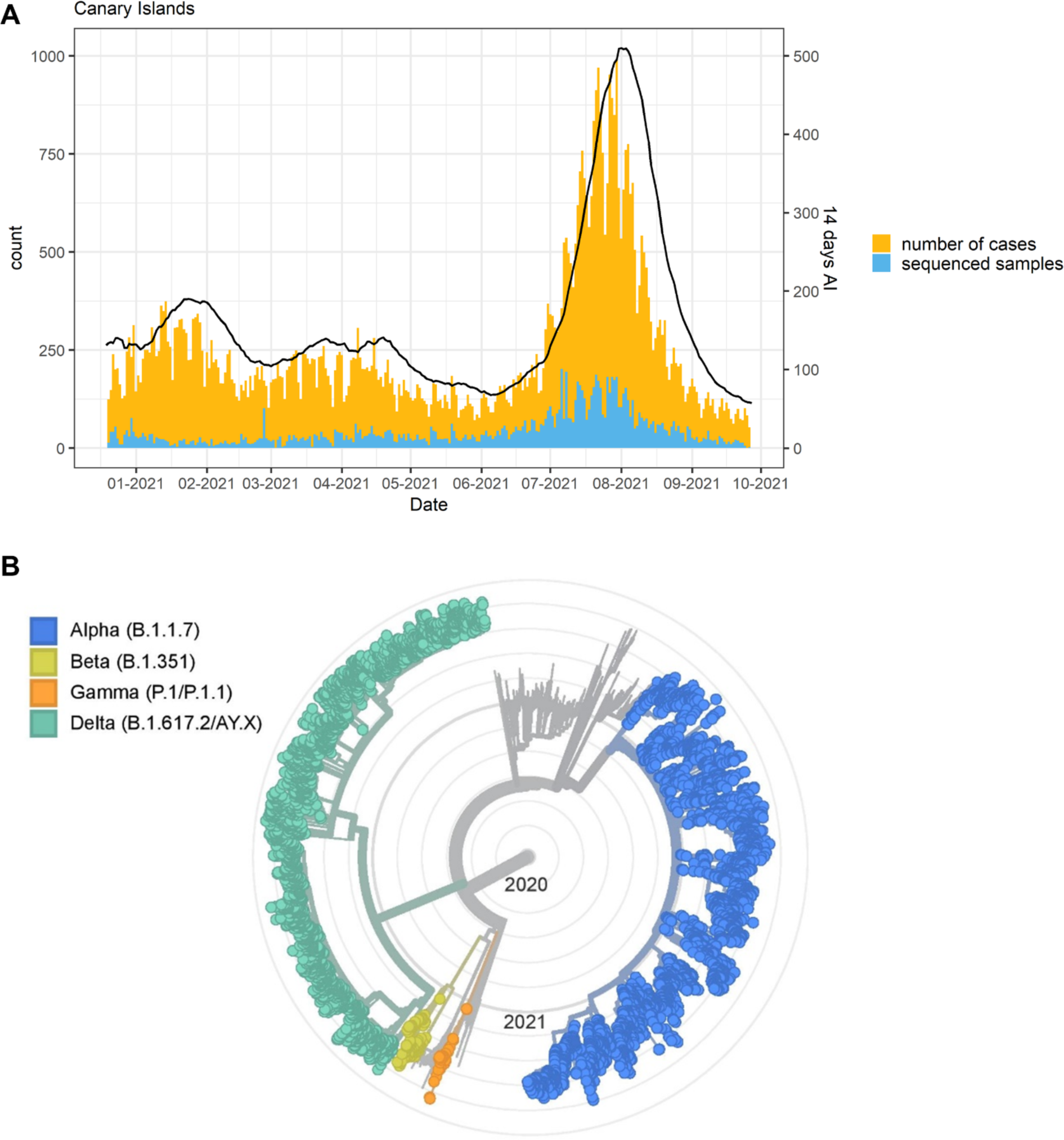
A) Number of cases, sequenced samples, and the 14-days accumulated incidence (AI, continuous line) throughout the study period in the Canary Islands archipelago. B) Phylogenetic tree (radial) showing the four VOCs with the background phylogenetic tree of samples collected in the Canary Islands throughout the study period.

The temporal distribution of the different VOCs in the Canary Islands is represented in **Figure 2**. Since there were relatively few samples sequenced from two of the smallest islands (La Gomera (n= 17) and El Hierro (n=50)) (**Figure S1**) and the samples from La Graciosa were collected within those from Lanzarote, we excluded these islands from the following description. As it was previously reported for Tenerife alone (Alcoba-Florez *et al*., 2021), the first description of Alpha in the Canary Islands occurred around the 23^rd^ of December 2020 and was associated with an increase in the 14-day accumulated incidence (**Figure 2A**). After that, the replacement of non-VOC lineages by the Alpha variant occurred more sharply in Gran Canaria and Lanzarote than in the other islands (Tenerife, Fuerteventura, and La Palma) with almost the totality of sequenced cases belonging to Alpha already by early February 2021 (**Figure 2B and 2C**).

**Figure 2.**
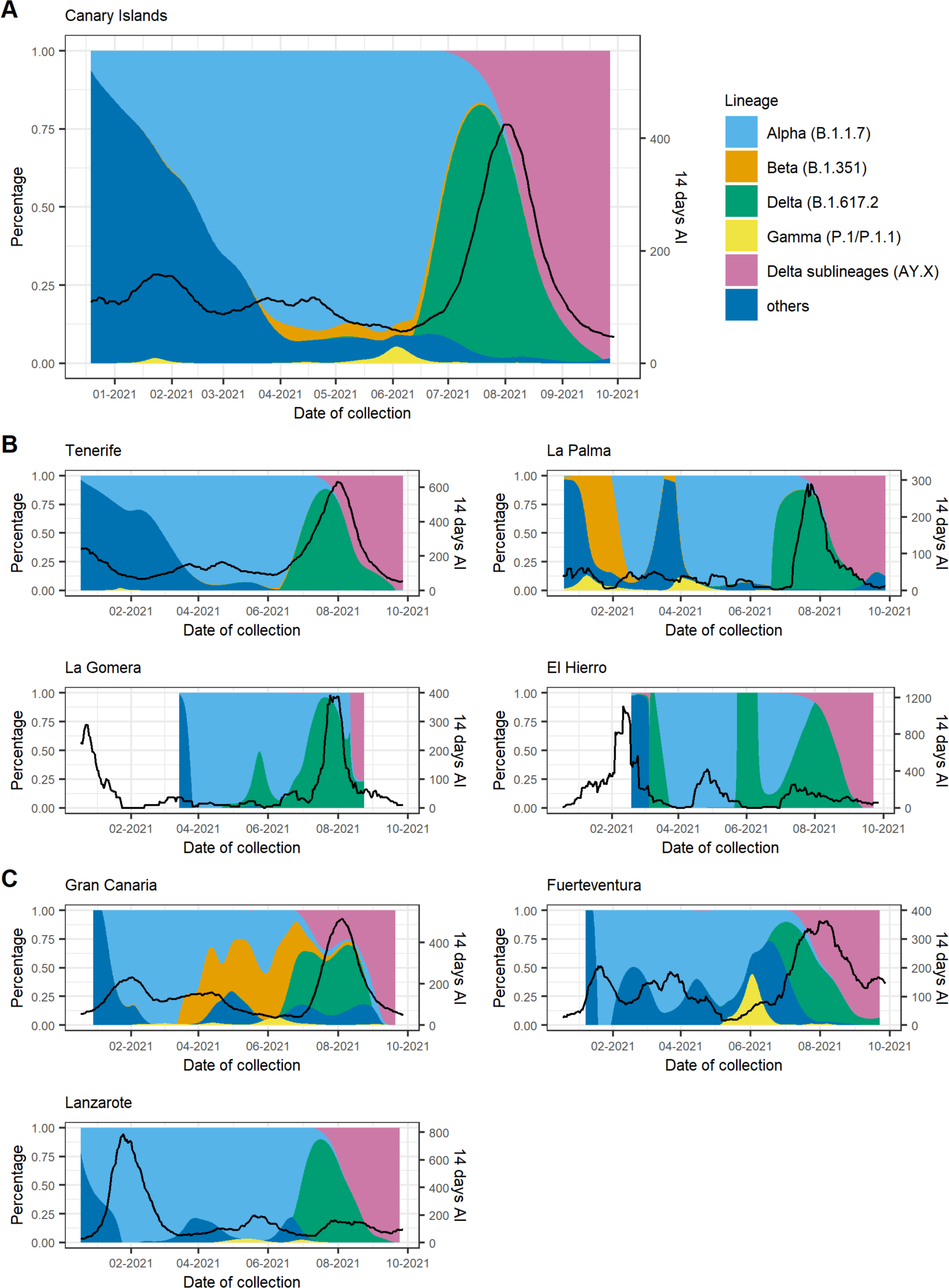
Proportion of VOCs and the 14-days accumulated incidence throughout the study period for A) the Canary Islands as a whole and disaggregated by island for B) the Tenerife province and C) the Gran Canaria province. Black lines depict the 14-day accumulated incidence. AY.X denotes the sub-lineages of Delta (B.1.617.2).

Apart from a first imported case in La Palma at the end of January 2021 due to a traveller returning from Cameroon, community transmission of Beta variant in the archipelago was first identified in the island of Gran Canaria, where a large cluster of 46 cases was observed between the end of March and mid-May 2021. At the beginning of April, a second small cluster of six cases was observed in Gran Canaria with no apparent connection to the first one. In Tenerife, three small clusters of an average of four cases were observed, one at the beginning of April and two throughout June. In mid-May, a second big cluster of 58 cases was observed in Gran Canaria which lasted up to the beginning of August, when the last case of this variant was observed. Almost no cases of the Beta variant were found on any other island throughout the study period. Cases of the Gamma variant were scarce and observed in Tenerife, Gran Canaria, La Palma, and in Lanzarote up to the end of May 2021, when a cluster of 24 P.1.1 cases was observed in Fuerteventura. The Fuerteventura outbreak was associated with a religious celebration involving local residents and foreign visitors and lasted up to the middle of June. An overview of the Canary Islands archipelago geography and of the heterogeneous distribution of Beta and Gamma variants in the archipelago is presented in **Figure 3**.

**Figure 3.**
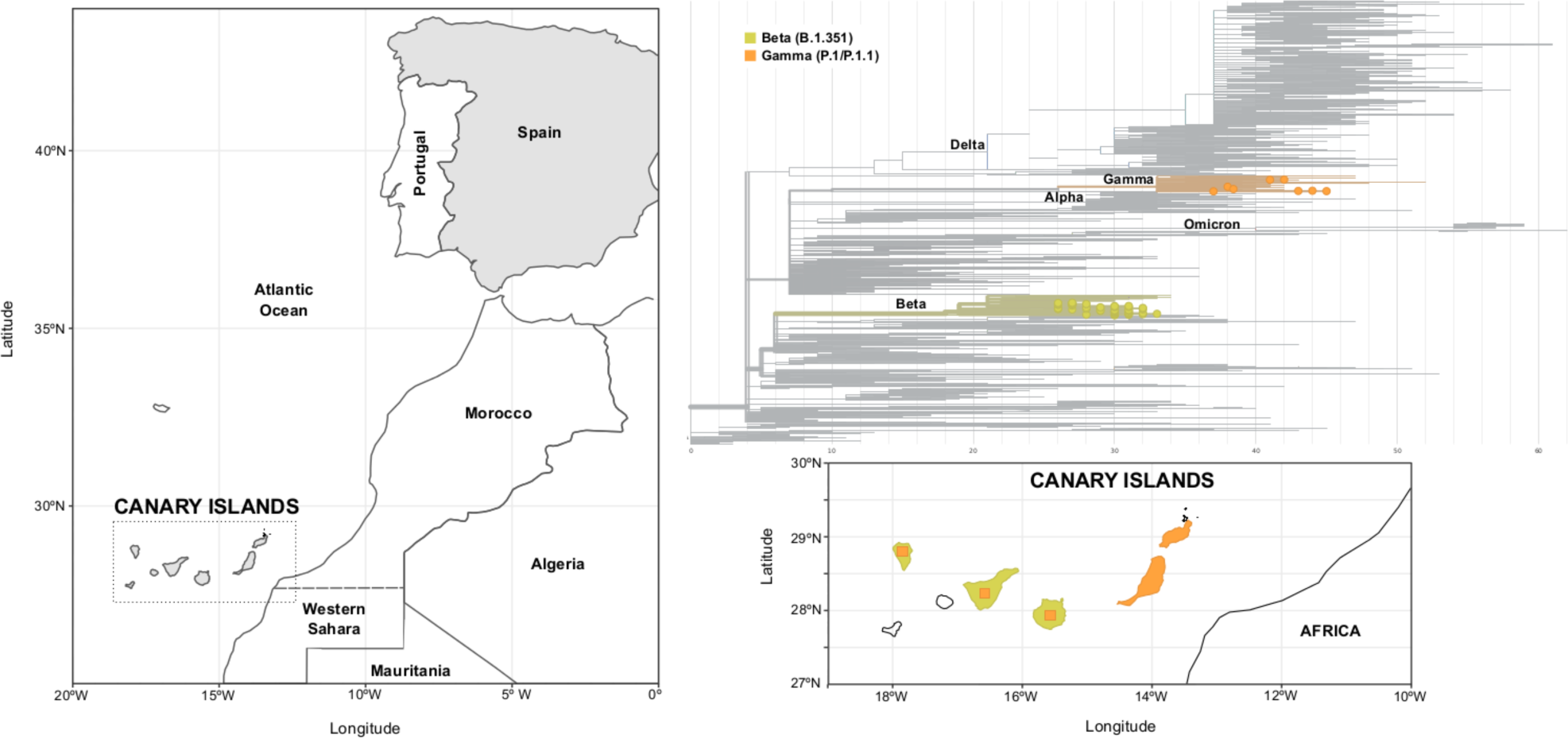
Geographical location of the Canary Islands, with respect to the Iberian Peninsula and Northwest Africa (left panel). Phylogenetic tree of Beta and Gamma variants (upper right panel) and their distribution in the islands of the archipelago (lower right panel). Square areas are proportional to the presence of the variant in each island.

Alpha, Beta, and Gamma lineages were replaced by the Delta variant which started circulating in Tenerife at the beginning of June 2021 (the first case detected in Tenerife on the 17^th^ of May) followed by its identification in all other islands, probably due to multiple introductions of this variant in the archipelago. The presence of Delta was associated with a rapid surge of COVID-19 cases in all islands when the archipelago entered the fifth wave and the highest number of cases was reached since the beginning of the pandemic. From the end of July 2021, the previous heterogeneous distribution of VOCs in the different islands converged to a similar figure, with Delta becoming prevalent and almost completely displacing all other variants circulating in the archipelago. At the beginning of August, the sub-lineages of Delta (mainly AY.4 and AY.12) started circulating in the islands, completely superseding Delta by the end of September 2021.

## Discussion

In this work, we dissected the spread of VOCs in the Canary Islands archipelago in the period between mid-December 2020 and late September 2021. We observed that the viral sequences detected in the islands were characterized by a diverse epidemiological background and variant distribution throughout the study, which can be largely explained by their geographical isolation and heterogeneous population size. While Alpha spread ubiquitously across the archipelago early after its first appearance in the island of Tenerife, Beta and Gamma variants were observed only in some of the islands and not in others. Beta was observed in the archipelago from late January up to August 2021, and its spatial distribution was mainly circumscribed to the islands of Gran Canaria and Tenerife, even though it was already present in several European countries by mid-January (O’Toole *et al*., 2021), representing up to 20% and 8% of cases in Austria and France, respectively, at that moment (Hodcroft, 2021). Cases associated with the Gamma variant were also relatively few, with a small cluster of cases observed in Tenerife in late January 2021, followed by isolated cases in the other islands. The largest Gamma cluster was a P.1.1 cluster observed in Fuerteventura. Both Beta and Gamma did not spread throughout the archipelago such as Alpha, something that was also observed in other European countries (Hodcroft, 2021). When Delta emerged, it was associated with a surge of cases in the archipelago throughout the summer, mimicking what occurred in the UK (Riley *et al*., 2021) and in the rest of Europe (Hodcroft, 2021). We attribute this not only to the intrinsically higher transmissibility of the Delta variant compared to the previously circulating VOCs (Campbell *et al*., 2021) but also to the concurrent lifting of COVID-19 restrictions in the archipelago and nationwide. Since August 2021, we observed the emergence of diverse Delta sub-lineages (AY.4 and AY.12), similar to what occurred worldwide throughout the summer (Angeletti *et al*., 2021; Eales *et al*., 2021).

In summary, as a response to the COVID-19 pandemic, we established a network for genomic surveillance of SARS-CoV-2 in the Canary Islands based on two sequencing technologies, leveraging homogeneous and centralized processing of samples, efficient sequencing workflows, rigorous data quality control, and accurate sequence classification. To do so, we relied on both previously existing capacities and human resources in the involved laboratories, and the rapid dissemination of information over VOCs and open-source bioinformatics tools made available by researchers worldwide. We managed to sequence more than 15% of COVID-19 cases of the archipelago in the study period, aligning our sequencing strategy to the WHO and the ECDC recommendations. We recognize sampling bias as a limitation of our study, with sample collection being not proportional to the number of cases in all islands. In fact, the island of Tenerife always had the highest percentage of sequenced samples, mainly because the network was physically located in the island and therefore sample logistics was easier. Despite this, we managed to efficiently identify and track all VOCs in the Canary Islands since December 2020 and promptly inform the Public Health authorities in the region.

## Data Availability

All data produced in the present study are available at GISAID.

## Acknowledgements

We deeply acknowledge the University Hospital Nuestra Señora de Candelaria and the Instituto Tecnológico y de Energías Renovables board of directors for their strong support and assistance in accessing diverse resources used in the study.

## Conflicts of interest

The authors declare that they have no known competing financial interests or personal relationships that could have appeared to influence the work reported in this paper.

## Funding

This research was funded by Cabildo Insular de Tenerife [grants CGIEU0000219140 and “Apuestas científicas del ITER para colaborar en la lucha contra la COVID-19”]; the agreement with Instituto Tecnológico y de Energías Renovables (ITER) to strengthen scientific and technological education, training research, development and innovation in Genomics, Personalized Medicine and Biotechnology [grant number OA17/008]; Ministerio de Ciencia e Innovación [grant number RTC-2017-6471-1], co-funded by the European Regional Development Fund (ERDF). The funders had no role in the study design, collection, analysis and interpretation of data, in the writing of the manuscript or in the decision to submit the manuscript for publication.

## Ethical Approval

The University Hospital Nuestra Señora de Candelaria (Santa Cruz de Tenerife, Spain) review board approved the study (ethics approval number: CHUNSC_2020_24).

## Supplementary Information

**Figure S1.**
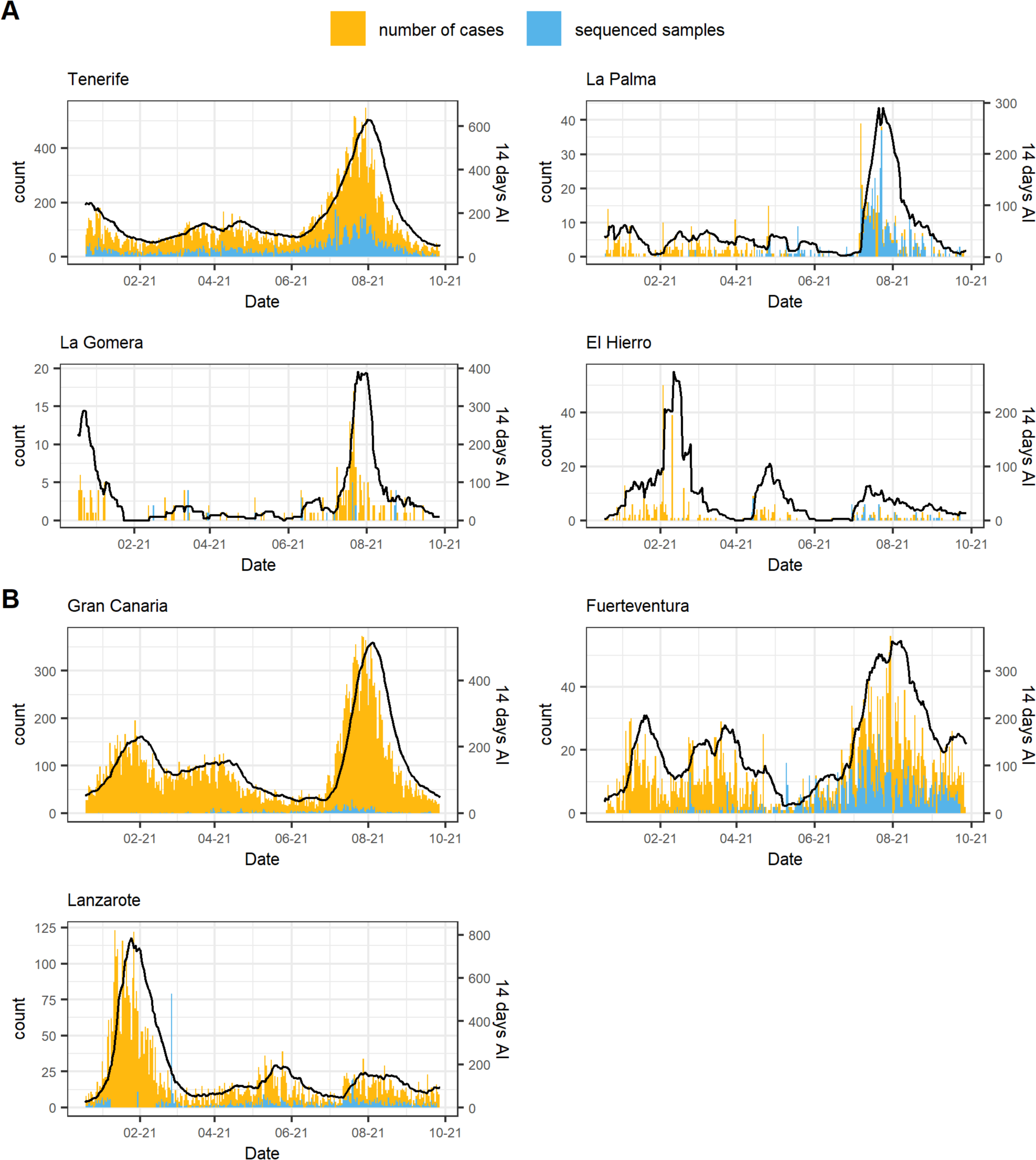
Number of cases, sequenced samples, and 14-days accumulated incidence (AI, continuous line) throughout the study period disaggregated by island (A) in the Tenerife province and B) in the Gran Canaria province.

## Notes

### Competing Interest Statement

The authors have declared no competing interest.

### Funding Statement

This research was funded by Cabildo Insular de Tenerife [grants CGIEU0000219140 and Apuestas cientificas del ITER para colaborar en la lucha contra la COVID-19]; the agreement with Instituto Tecnologico y de Energias Renovables (ITER) to strengthen scientific and technological education, training research, development and innovation in Genomics, Personalized Medicine and Biotechnology [grant number OA17/008]; Ministerio de Ciencia e Innovacion [grant number RTC-2017-6471-1], co-funded by the European Regional Development Fund (ERDF). The funders had no role in the study design, collection, analysis and interpretation of data, in the writing of the manuscript or in the decision to submit the manuscript for publication.

### Author Declarations

The University Hospital Nuestra Señora de Candelaria (Santa Cruz de Tenerife, Spain) review board approved the study (ethics approval number: CHUNSC_2020_24).

